# Economic Evaluation of an Employee Health Screening Program for Hepatitis C Virus in Pakistan

**DOI:** 10.1101/2025.08.28.25334700

**Authors:** Asra Qureshi, Theodore Bolas, Unab Inayat Khan, Ashar Muhammad Malik, Nicholas Risko

## Abstract

**Background:** Hepatitis C virus (HCV) causes significant global morbidity and mortality, and combating it is a global health priority. Pakistan has a particularly high burden of HCV, including cases that remain undetected and untreated.

**Methods:** In 2019, a pilot workplace health screening program for chronic diseases, including HCV, was launched at Aga Khan University in Pakistan. We performed an economic evaluation to determine its cost-effectiveness in screening for HCV, inform resource allocation decisions, and advance evidence in this field. Over the course of the screening pilot, 1,809 employees completed enrollment, blood tests, assessment, and consultation. Cost-effectiveness modeling was conducted using a combination of decision-tree and Markov approaches. Univariate and multivariate probabilistic sensitivity analyses were performed.

**Results:** The median final ICER was $307.39 per DALY averted. Uncertainty around the health status of those lost to follow-up produced a large confidence interval, from $167.92 to $1601.22 per DALY averted, depending on the assumptions made. This screening is estimated to save anywhere from 1 to 14 lives by prevention of HCV-induced liver failure, depending on the burden of HCV in the lost to follow-up group, out of a total of 25 HCV-induced liver failure deaths in the control group, between a 44% and a 4% reduction.

**Conclusions:** Our findings support that employee screening for HCV is cost-effective in high-prevalence areas such as Pakistan and a wise investment of health resources. The findings also point to programmatic improvements that could maximize the health benefits and improve the cost-effectiveness of other similar programs.

## Background

Hepatitis C virus (HCV) is a bloodborne virus that causes inflammation of the liver, causing the disease Hepatitis C, which has both acute and chronic stages. Globally, an estimated 58 million people have chronic infections, with about 1.5 million new infections occurring per year. (1) Chronic HCV infection leads to liver fibrosis, cirrhosis, and hepatocellular carcinoma (HCC), leading to more than 350,000 deaths annually. (2, 3) Advances in HCV treatment have led to a commitment from all 194 member states of WHO to eliminate viral hepatitis as a public health threat. (4)

Pakistan currently has one of the highest prevalence rates globally (≈5-6%), with an estimated 8-11 million individuals infected with HCV. (5, 6) Moreover, there is a large burden of undiagnosed infections in the country. While generic direct-acting antivirals (DAAS) are available in Pakistan at a price as low as USD 60 per treatment course, a lack of population-based screening and awareness hinders elimination rates. (5) Until recently, no formal steps on population-based screening for HCV have been taken. In 2017, the Ministry of Health-Government of Pakistan launched its first National Hepatitis Strategic Framework (NHSF) (7) to offer access to safe, affordable, and effective prevention, care, and treatment services for viral hepatitis. However, there continue to be critical gaps and resource limitations around screening and linkage to treatment. (5) Furthermore, studies have shown that in general, people have inadequate health literacy about Hepatitis C, including a lack of basic knowledge of the transmission mode, and other misconceptions about the disease. (8) (9)

In 2019, Aga Khan University in Pakistan piloted an Employee Health and Wellness Program (EHWP) that screened full-time employees for common chronic diseases such as anemia, hypertension, diabetes, dyslipidemia, depression, and HCV. As routine preventive health checks are not the norm in Pakistan and HCV testing services have historically only been offered at tertiary care centers, it was felt that on-the-job screening could offer a solution. A significant rise in local prevalence over recent years made this a pressing issue. (5, 6, 10, 11) Details about program structure, execution, methodology, evaluation, and initial findings have already been published and can be accessed at Khan UI et.al. (12)

Economic considerations maintain a prominent role in the planning, management, and evaluation of health systems. (13) To examine the costs and outcomes of HCV screening, with an aim to inform program development and policy in this realm we conducted an economic evaluation.

## Methods

The full-time employees who were screened obtained free laboratory testing, received an individual health risk assessment, and if required were referred for further treatment or behavioral counseling. Informational sessions were conducted across various departments of the University. Details of employees who completed the program and Hepatitis C costing details are shown in Tables 1 and 2 respectively.

**Table 1.**
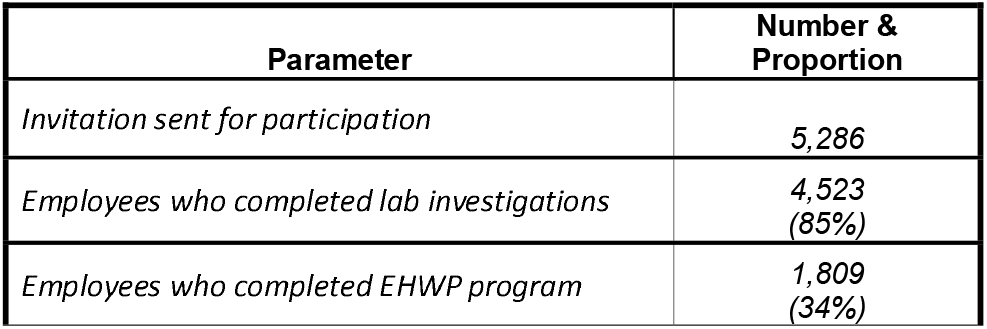
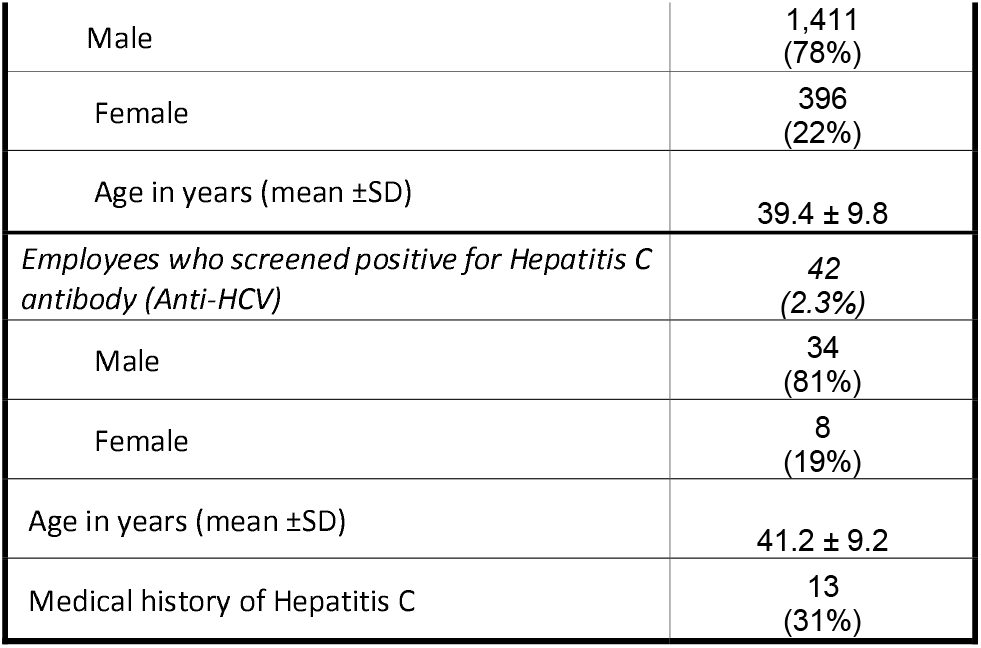
Characteristics of employees who completed risk assessments and Hepatitis C screening status.

**Table 2.** Costing incurred on Hepatitis C screening in EHWP.

| Parameter | Cost | Reference |
| --- | --- | --- |
| Anti HCV test cost | 2400 PKR<br>17.1 USD | Cost assigned for HCV testing under EHWP |
| Nursing assessment cost | 150 PKR<br>1.07 USD | Cost assigned for HCV testing under EHWP |
| HCV Treatment Cost | 8,421 PKR<br>60 USD | (5) |

### a. Analytic and modeling approach

We performed a cost-effectiveness analysis of the HCV arm of the pilot, to estimate the costs and benefits of screening employees for HCV. Benefits were measured in terms of new HCV cases identified and Disability Adjusted Life Years (DALYs) averted. We consider lifetime costs and benefits incurred to the employees who completed the program as the time horizon for economic analysis. All analysis was conducted in R and RStudio. (14)

We took a program-level costing perspective, analyzing the costs to the employer to establish, and run the screening program. These costs include the cost of HCV testing, as well as assessment after testing. Costs were measured in 2019 local currency and adjusted to 2023 for the analysis, with conversion between time-matched international currencies where these inputs were needed. Costs to the employer do not include the loss of productivity suffered by the employer when their employees fall ill and thus miss work.

All costs and DALYs were discounted at a rate of 3% annually. Discounting is used in cost-effectiveness analysis studies to properly weigh immediate and long-term benefits and costs, based on the assumption that future benefits and costs are less valuable and less costly than those incurred in the present. The appropriate discount rate can vary based on the types of outcomes and societal values. Most global health studies use a rate of 3% annually, which was chosen for this study to ensure comparability across papers. (13, 15)

To estimate the benefits of HCV screening, we simulated the health progressions of the original cohort of employees, using a decision tree and Markov model. For simplicity, the decision tree shows only employees who are Hep-C positive, as those who are negative will end up in the same health state as the Markov model (not infected.) The decision tree shows the probability of infected employees starting the Markov model either in sustained virological recovery (SVR) or F0 stage of HCV. The Markov model uses established HCV data to simulate the cohort’s health trajectories over their lifespan, comparing the DALYs (disability-adjusted life years) accumulated in the presence and absence of the program.

The decision tree initially branches into the standard of care and program arms (Figure 1). From there, HCV-positive patients have a chance of being tested, and if tested, a probability of being treated, and if treated, a probability of entering sustained virological recovery (SVR). In the standard routine care, patients have an equal probability of having HCV but have a decreased likelihood of being tested, and a similar likelihood of being treated conditional on being tested. In the treatment group, the likelihood of testing increases, leading to a greater number of treated patients entering the Markov model in SVR.

**Figure 1.**
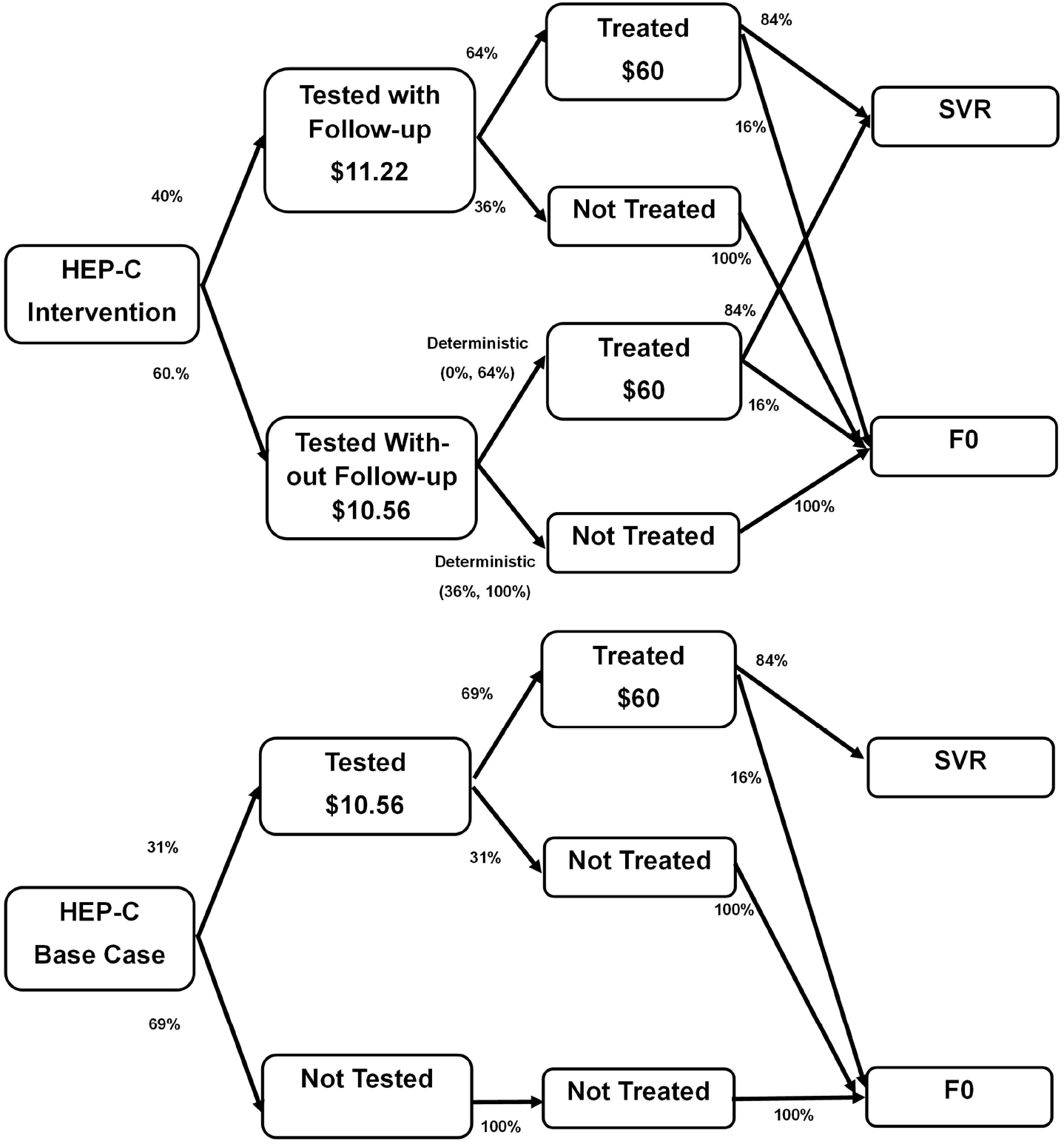
Decision tree

The probability of being tested and treated in the presence of the program is taken from EHWP data. The probability of being tested and treated outside of the program is taken from the number of people with an existing HCV diagnosis, who independently sought treatment before the start of the program. The prevalence of HCV in our study population is estimated from the prevalence of positive cases among those who were tested and attended a screening appointment. Probability of SVR comes from a previous study of the cost-effectiveness of HCV treatment in Pakistan. (16)

Our Markov model (Figure 2) is derived from a published cost-effectiveness study on the use of direct-acting antivirals for treating HCV in Pakistan (16), as it has been previously validated and is specific to the Pakistani context. Patients with HCV can progress linearly along the Metavir HCV stages (F0-F4). Because our patients were not staged when tested, they were all assumed to start in the F0 stage, the most conservative assumption in terms of cost-effectiveness. Because our program did not track patients after treatments began, the original model was simplified from an infected to under-treatment to treated model to one where patients were either infected or in sustained virological recovery. Patients in the F4 stage of compensated cirrhosis can pass to decompensated cirrhosis (DC), and those in either stage of cirrhosis can develop hepatocellular carcinoma (HCC). As in the cited model, all patients face an annual death rate taken from Pakistani life tables (17), which are dependent on the average age of the cohort. Patients in DC or HCC states also face an additional death rate from HCV. All patients with HCV are assumed to start in the F0 stage, and patients can progress through 1 health state each year.

**Figure 2.**
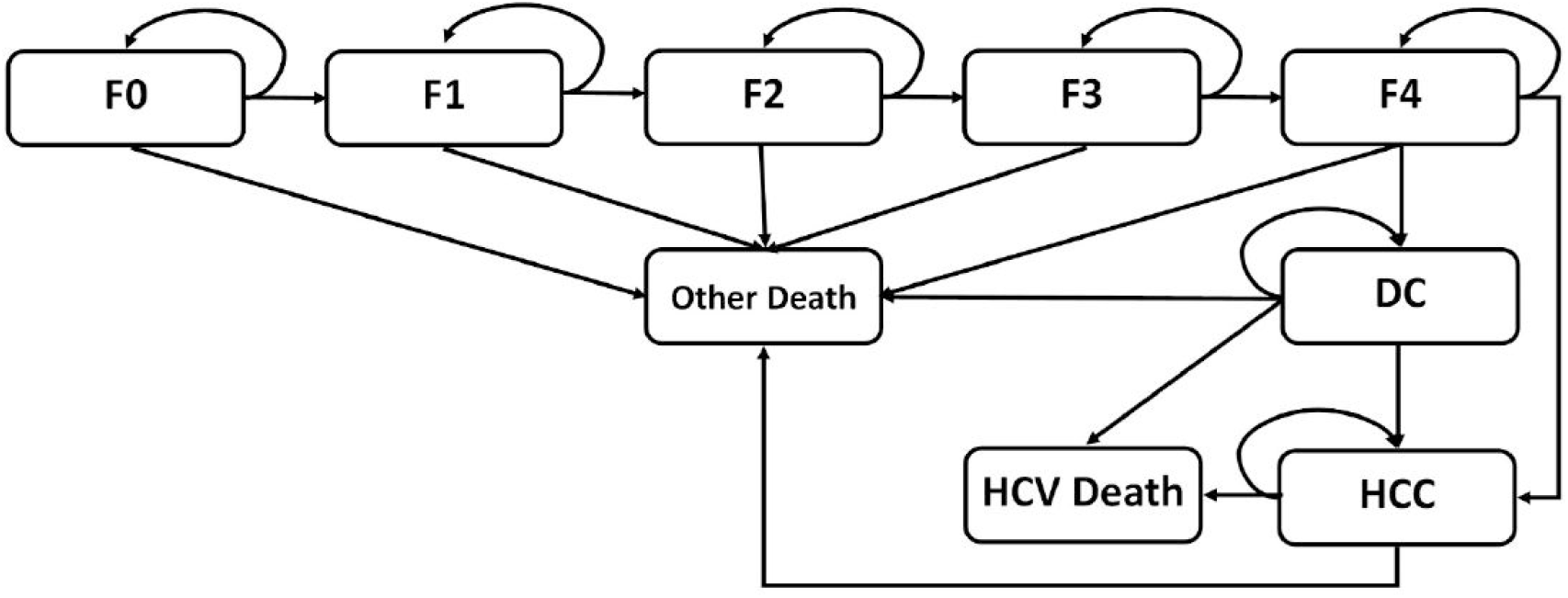
Markov Model

Each person’s year spent in a health state is multiplied by that health state’s corresponding disability rate to calculate the total DALYs accumulated for a given year. Disability rates for F4, DC, and HCC states were taken from the Global Burden of Disease study (18), and disability rates for mild to moderate HCV were calculated as percentages of the F4 state following the precedent of published literature. (16, 17) Recovered and never-infected DALYs are assumed to be zero, while death is given a DALY weight of 1. The model was allowed to run for 46 years until the cohort age was 85, the last age with data on Pakistani life tables.

While the program did not provide treatment directly, cost of treatment was included in the model, based on prices for direct-acting antivirals. (5) This does not include potential costs associated with untreated Hepatitis C, as data are not reliably available in the Pakistani context. While including these costs might capture an even stronger benefit of this program for the healthcare system, given the weakness of evidence, and the fact that their omission biased our results towards the null, we chose to omit these.

### b. Sensitivity Analysis

We used both a scenario analysis and a probabilistic sensitivity analysis to examine the robustness of our results. In our scenario analysis, we varied only 1 parameter, the odds that a tested patient who was lost to follow-up received treatment. In one scenario, we model the odds of treatment at 0, and in the other scenario, the odds of treatment are the same as those who were not lost to follow-up. This variable was selected to explore the vulnerability of the model to this key parameter given the high number of participants lost to follow-up in the initial study and the potential to mitigate this in future implementation efforts. The reason why patients who were lost to follow-up could still receive treatment is that the treatment for all patients was done outside of the program by other healthcare providers, so they would have been likely to seek outside care if they received a positive result.

We also performed a multivariate probabilistic sensitivity analysis, allowing the relevant epidemiological parameters to vary as a normal, binomial, uniform, or beta distribution where appropriate. These probability distributions were drawn independently 1,000 times, to generate 1000 different ICER values. All parameters, as well as their distribution parameters, are shown in Table 3. All distribution parameters came from their respective paper which reported the means. Program costs were not allowed to vary as exact costs were known for this project, but treatment costs varied as a gamma distribution. All parameters varied independently of one another.

**Table 3.** Sensitivity Analysis Parameters.

| Parameter | Deterministic Value | Distribution Parameter | Distribution Used |
| --- | --- | --- | --- |
| F0-F1 Transition<sup>*</sup> | 0.117 | $\mu=0.117, \sigma=0.0051$ | Normal |
| F1-F2 Transition<sup>*</sup> | 0.085 | $\mu=0.085, \sigma=0.0036$ | Normal |
| F2-F3 Transition<sup>*</sup> | 0.121 | $\mu=0.121, \sigma=0.0046$ | Normal |
| F3-F4 Transition<sup>*</sup> | 0.115 | $\mu=0.023, \sigma=0.0041$ | Normal |
| F4-DC Transition<sup>*</sup> | 0.039 | (14.6,360.2) | Beta |
| F4-HCC Transition<sup>*</sup> | 0.14 | (1.9,11.61) | Beta |
| DC-HCC Transition<sup>*</sup> | 0.14 | (1.9,11.61) | Beta |
| DC-HCV Death Transition<sup>*</sup> | 0.13 | (0.11,0.15) | Uniform |
| HCC-HCV Death Transition<sup>*</sup> | 0.43 | (0.37,0.49) | Uniform |
| DALY HCV Stage F1 | 0.25*F4 | -- | -- |
| DALY HCV Stage F2 | 0.5*F4 | -- | -- |
| DALY HCV Stage F3 | 0.75*F4 | -- | -- |
| DALY HCV Stage F4 | 0.114 | (0.078, 0.159) | Uniform |
| DALY DC | 0.194 | (0.123, 0.250) | Uniform |
| DALY HCC | 0.451 | (0.307, 0.6) | Uniform |
| Proportion of Infected in SVR * | 0.84 | $p=0.84, n=1952$ | Binomial |
| Prevalence of Hepatitis C | 0.023 | $\mu=0.023, \sigma=0.0023$ | Normal |
| Treatment Rate in those lost<br>to follow-up ** | 0 & 0.64 | (0, 0.64) | Uniform |
| Cost of Treatment | 60 | $\alpha = 600, \beta = 0.1$ | Gamma |
| *Distribution parameters are as follows: Normal parameter is standard deviation. Beta parameter is (Alpha, Beta). Uniform parameter is (Low, High). Binomial parameter is population size.*<br>*Source: \*Mafirakureva et. Al cost-effectiveness of screening and treatment using direct acting anti virals (5)*<br>*\*\*EHWP program data* |  |  |  |

All uncertain parameters were chosen to vary, and a standard deviation of 10% was used if unknown. This allowed for some robustness in the model to defects in existing data, and changes in parameters over time.

## Results

The scenario analysis found that this program averts between 30.74 and 307.64 DALYs over the lifespan of the participants, on average saving between 0.066-0.007 DALYs per participant. At a total cost of $48,956.82 the Incremental Cost Effectiveness Ratio (ICER) is between $159.13, and $1592.45 spent per DALY averted as shown in Table 4. This specific program was estimated to save anywhere from 1 to 14 lives by prevention of HCV-induced liver failure, depending on the burden of HCV in the lost to follow-up group, out of a total of 25 HCV-induced liver failure deaths in the control group, between a 4% and a 44% reduction. Costs are the same in all groups, as the payer perspective is that of the university, which only paid for testing and follow-up. Treatment costs, which would vary between these two scenarios, are out of the scope of this model.

**Table 4.** Scenario Analysis Results.

| Parameter | Total Costs | DALYs | ICER | HCV Related Deaths |
| --- | --- | --- | --- | --- |
| Standard of Care | \$1,350 | 29266 | | 35 |
| Program (Low Treatment in group lost to follow-up scenario) | \$50,577 | 29235 | | 34 |
| Program (High Treatment in group lost to follow-up scenario) | \$ 53,007 | 28958 | | 21 |
| Incremental (Low Treatment in group lost to follow-up scenario) | \$49,227 | 31<br>(Averted) | 1601 | 1<br>(Averted) |
| Incremental (High Treatment in group lost to follow-up scenario) | \$51,657 | 308<br>(Averted) | 168 | 14<br>(Averted) |

Refined with the multivariate probabilistic sensitivity analysis, our model produced a final median ICER value of $307.39 per DALY averted, with a distribution ranging between $167.92-1601.22 per DALY averted (Figure 3).

**Figure 3.**
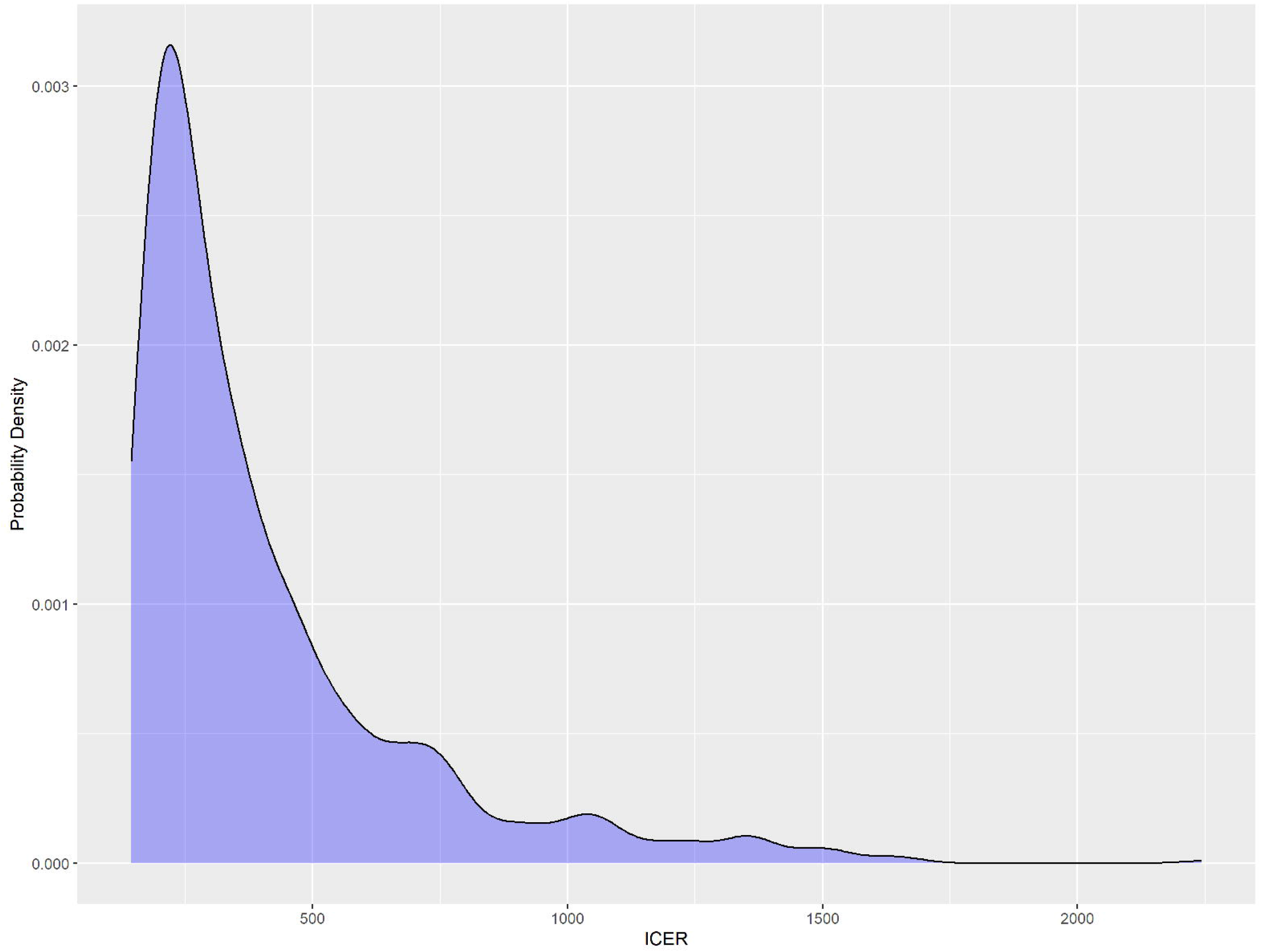
Probability Distribution of ICER over 1,000 simulations with variable parameters

## Discussion

Treating HCV-related infections at an early-stage results in halting the progression to advanced clinical stages that require higher treatment costs and lead to higher morbidity and mortality. (16) In developing countries like Pakistan, where out-of-pocket (OOP) expenditure accounts for 67% of total expenditure on healthcare, (19) it is crucial to use the available resources sustainably. To date, very few studies have assessed the cost-effectiveness of similar programs, and we were unable to identify any studies specific to this setting and population.

Our study demonstrates that HCV screening offered through a workplace health screening program to employees, is economically effective for early identification and linkage to care in low-resource settings. In addition, it has the potential to avert significant morbidity and mortality. As noted above, by screening for Hepatitis C, our median final ICER was $307.39 spent per DALY averted. Issues in the pilot around loss to follow-up led to a wide confidence interval and we demonstrated in our sensitivity analysis the important health and economic benefits of improving tracking and decreasing attrition from the program.

Our final ICER is significantly lower than Pakistan’s per capita GDP of $1505.01. (20) Although cost-effectiveness thresholds remain controversial, WHO recommends that programs that spend less than three times the per capita GDP are considered “cost-effective” and those that spend one times the per capita GDP are considered “highly cost-effective”. (21) Programs that meet these standards should be prioritized for health funding.

An important strength of this study is the use of real data of employees who were tested, screened, and assessed as part of the ongoing EHWP, including the true costs of running the program. Our findings lend support to the position that HCV screening should be part of an employee’s health check-up in geographies with higher HCV prevalence rates and that such initiatives not only offer benefits to both employees and employers but are also a worthwhile use of resources from a public health perspective.

A study conducted in a primary care setting for Hep C screening and treatment in Pakistan reported an ICER of $572 per DALY (USD 2023). (16) This suggests that EHWP is performing similarly to existing highly cost-effective programs in the country and may be highly suitable for wider scale-up. Outside the employee health modality, a small amount of research has assessed screening programs, in other low-resource settings such as Egypt (21, 22) and India (23), with findings supporting such interventions as cost-effective.

### Limitations

As we did not have access to the results of employees lost to follow-up in the original study, we assumed that those who received their test results but did not complete the assessment were equally likely to have Hep C as those who continued in the program. This was a conservative assumption, as it is likely that those with a negative result were less likely to follow up. This assumption was explored in our sensitivity analysis and produced the wide confidence interval noted in our results. If the screening program missed a significant number of HCV cases due to attrition it would have lower cost-effectiveness, and vice versa. We believe the median ICER derived from our probabilistic analysis most likely represents the true cost-effectiveness of the EHWP. However, our recommendation to the EHWP and similar programs is to utilize programmatic tools to avoid such high attrition in the future, which will enhance the overall health impact and cost-effectiveness of the screening program.

Another limitation exists with regards to cost calculations, as costs to the employer do not include productivity loss from employees suffering from HCV, due to a lack of data in the Pakistani context. By excluding this element which would provide financial benefits weighted on the side of the intervention, the model increases the estimated net cost to the employer and provides more conservative results.

Finally, there are the inherent and universal limitations of Markov modeling and cost-effectiveness analyses. For example, we had to make a series of assumptions and simplifications on the progression and disability levels of HCV in Pakistan. Although we followed an existing model and the best available literature, there is still a shortage of data on the disability rate at each progressive stage of HCV. Because our model makes predictions about the outcomes of those infected based on existing epidemiological data rather than observing their actual outcomes, it can serve only as an estimate.

## Conclusions

According to our analysis, an employee health screening program for HCV in the high prevalence area of Karachi, Pakistan had a cost-effectiveness of $307.39 per DALY averted. This would categorize the program as highly cost-effective and suggest that similar programs be prioritized for health funding. The findings of our sensitivity analysis point towards the large potential benefits in health and cost-effectiveness of programmatic improvements that would diminish loss to follow-up of participants. Further research is needed on the costs and impacts of employee health screening in low-resource settings to inform decision-makers and policy.

## Data Availability

All data produced in the present study are available upon reasonable request to the authors

## Abbreviations

EHWP: Employee Health and Wellness Program
HCV: Hepatitis C Virus
OOP: Out of pocket expenditure
ICER: Incremental cost-effectiveness ratio

## Declarations

### a. Ethics approval and consent to participate

Ethical approval was obtained from Aga Khan University ethical review committee (#2019-1281-3520). Written informed consent was obtained from the participants before the commencement of the study and as per ERC requirement, a signed copy of the consent form was also provided to the participants for their record. All study procedures were performed in accordance with the Declaration of Helsinki

### b. Consent for publication

Not applicable

### c. Availability of data and materials

The dataset used for analysis of this research study can be made available from the corresponding author on reasonable request.

## Authors’ Contribution

AQ-primary author, contributed to the conceptualization, data curation, and team collaboration. AQ and TB wrote the manuscript (background, methods, results, and discussion). TB performed formal analysis and modeling, and prepared figures. UIK clinical expert provided clinical insight into the concept, and manuscript review and contributed to writing methods and discussion. AM contributed to data synthesis and manuscript review. NR-senior author contributed to research design, data analysis, modeling review, work supervision, and discussion writing. NR & AQ reviewed and formatted the manuscript. All authors have read and approved the submitted version of the manuscript.

## Acknowledgments

The authors are very thankful for the initial intellectual contribution of Dr Kalin Werner, PhD, who helped to conceive the study. We also would like to acknowledge the administrative and logistical support provided by the Human Resource Division; Information Technology Support Services; and the Employee Health team of Aga Khan University, Karachi, Pakistan.

## References

1. Hepatitis C: WHO; © 2022 WHO [Available from: https://www.who.int/news-room/fact-sheets/detail/hepatitis-c#:∼:text=Globally%2C%20an%20estimated%2058%20million,new%20infections%20occurring%20per%20year.

2. Dore G, Ward J, Thursz M. Hepatitis C disease burden and strategies to manage the burden (Guest Editors Mark Thursz, Gregory Dore and John Ward). Journal of viral hepatitis. 2014;21:1–4.

3. Averhoff FM, Glass N, Holtzman D. Global burden of hepatitis C: considerations for healthcare providers in the United States. Clinical infectious diseases. 2012;55(Suppl_1):S10–S5.

4. Heffernan A, Cooke GS, Nayagam S, Thursz M, Hallett TB. Scaling up prevention and treatment towards the elimination of hepatitis C: a global mathematical model. The Lancet. 2019;393(10178):1319–29.

5. Chhatwal J, Chen Q, Wang X, Ayer T, Zhuo Y, Janjua NZ, et al. Assessment of the feasibility and cost of hepatitis C elimination in Pakistan. JAMA network open. 2019;2(5):e193613–e.

6. Haqqi A, Munir R, Khalid M, Khurram M, Zaid M, Ali M, et al. Prevalence of hepatitis C virus genotypes in Pakistan: current scenario and review of literature. Viral immunology. 2019;32(9):402–13.

7. Organization WH. Address by Dr Mahmoud Fikri Regional Director WHO Eastern Mediterranean Region on the occasion of launch of the national hepatitis strategic framework 2017-2021, Islamabad, Pakistan, 8 October 2017. 2017.

8. Maqsood S, Iqbal S, Zakar R, Zakar MZ, Fischer F. Determinants of overall knowledge and health behaviours in relation to hepatitis B and C among ever-married women in Pakistan: evidence based on Demographic and Health Survey 2017–18. BMC Public Health. 2021;21(1):2328.

9. Younus I, Sarwar S, Qaadir A, Butt Z, Nazir A, Choudry A. Correlation of literacy and awareness regarding hepatitis C: a survey of family members of hepatitis C patients. Int J Med Res Rev. 2016;4(09):1576–81.

10. Khalid GG, Kyaw KWY, Bousquet C, Auat R, Donchuk D, Trickey A, et al. From risk to care: the hepatitis C screening and diagnostic cascade in a primary health care clinic in Karachi, Pakistan—a cohort study. International Health. 2020;12(1):19–27.

11. Qureshi H, Bile K, Jooma R, Alam S, Afrid H. Prevalence of hepatitis B and C viral infections in Pakistan: findings of a national survey appealing for effective prevention and control measures. EMHJ-Eastern Mediterranean Health Journal, 16 (Supp), 15–23, 2010. 2010.

12. Khan UI, Qureshi A, Lal K, Ali S, Barkatali A, Nayani S. Implementation and evaluation of Employee Health and Wellness Program using RE-AIM framework. International Journal of Workplace Health Management. 2021.

13. Cost-Effectiveness Analysis: CDC-Office of the Associate Director for Policy and Strategy; [updated October 20, 2021. Available from: https://www.cdc.gov/policy/polaris/economics/cost-effectiveness/index.html#:∼:text=CEA%20can%20be%20useful%20in,compared%20to%20an%20alternative%20intervention.

14. R: A language and environment for statistical computing. R Foundation for Statistical Computing, Vienna, Austria. 2021.

15. WHO-Life tables Mortality and Global health data repository-life tables by country: WHO; 2020 [Available from: https://apps.who.int/gho/data/view.main.61230?lang=en.

16. Mafirakureva N, Lim AG, Khalid GG, Aslam K, Campbell L, Zahid H, et al. Cost-effectiveness of screening and treatment using direct-acting antivirals for chronic Hepatitis C virus in a primary care setting in Karachi, Pakistan. Journal of viral hepatitis. 2021;28(2):268–78.

17. Mafirakureva N, Dzingirai B, Postma MJ, van Hulst M, Khoza S. Health-related quality of life in HIV/AIDS patients on antiretroviral therapy at a tertiary care facility in Zimbabwe. AIDS care. 2016;28(7):904–12.

18. Global Burden of Disease (GBD) [English]. IHME-THE INSTITUTE FOR HEALTH METRICS AND EVALUATION (University of Washington); 2019 [Available from: https://www.healthdata.org/research-analysis/gbd.

19. Muhammad Malik A, Azam Syed SI. Socio-economic determinants of household out-of-pocket payments on healthcare in Pakistan. International journal for equity in health. 2012;11(1):1–7.

20. GDP per capita (current US$) - Pakistan: The World Bank; 2023 [Available from: https://data.worldbank.org/indicator/NY.GDP.PCAP.CD?locations=PK.

21. Griffiths M, Maruszczak M, Kusel J. The who-choice cost-effectiveness threshold: a country-level analysis of changes over time. Value in Health. 2015;18(3):A88.

22. Obach D, Deuffic-Burban S, Esmat G, Anwar WA, Dewedar S, Canva V, et al. Effectiveness and cost-effectiveness of immediate versus delayed treatment of hepatitis c virus–infected patients in a country with limited resources: The case of egypt. Clinical infectious diseases. 2014;58(8):1064–71.

23. Aggarwal R, Chen Q, Goel A, Seguy N, Pendse R, Ayer T, et al. Cost-effectiveness of hepatitis C treatment using generic direct-acting antivirals available in India. PloS one. 2017;12(5):e0176503.

